# In-Hospital Mortality Trends Among Hospitalized Adults With Cardiac Arrest, 2016-2022

**DOI:** 10.64898/2026.09.25.26364059

**Authors:** Rahul Nedunuri, Mehrtash Hashemzadeh, Mohammad Reza Movahed

**Affiliations:** University of Arizona College of Medicine–Phoenix; University of Arizona Sarver Heart Center, Tucson

**Keywords:** In-Hospital Cardiac Arrest (IHCA), Out-of-Hospital Cardiac Arrest (OHCA), Cardiopulmonary Resuscitation (CPR), Chest Compression-Only CPR (CCO-CPR), Resuscitation, Return of Spontaneous Circulation (ROSC)

## Abstract

**Background:** CPR is the primary intervention for cardiac arrest, and the mortality trends before and after the pandemic remain are not fully understood.

**Methods:** This cross-sectional study used the National Inpatient Sample (NIS) database from 2016-2022 and International Classification of Diseases, Tenth Revision (ICD-10) codes to evaluate the mortality trend for adult patients undergoing CPR after in-hospital cardiac arrest (IHCA). Primary exposures were calendar year, demographics, socioeconomic factors, and hospital characteristics. Multivariable weighted logistic regression was performed to identify predictors of mortality after CPR for IHCA.

**Results:** Among 612,930 weighted adult hospitalizations undergoing CPR for IHCA between 2016 and 2022, overall in-hospital mortality was 67.8%. Risk-adjusted mortality declined through 2019 before increasing sharply during the COVID-19 pandemic, peaking in 2021 and remaining above pre-pandemic levels in 2022. Compared with 2016, admission in 2020 (aOR 1.18, 95% CI 1.12–1.25; p<0.001) and 2021 (aOR 1.26, 95% CI 1.19–1.33; p<0.001) was independently associated with higher mortality, whereas admission in 2019 was associated with lower mortality (aOR 0.89, 95% CI 0.84–0.95; p<0.001). Higher mortality was also associated with increasing age (aOR 1.01 per year; p<0.001), female sex (aOR 1.05, 95% CI 1.02–1.08; p<0.001), Black (aOR 1.14, 95% CI 1.10–1.18; p<0.001) and Hispanic race/ethnicity (aOR 1.20, 95% CI 1.15–1.26; p<0.001), and self-pay insurance (aOR 1.62, 95% CI 1.51–1.74; p<0.001). Protective factors included private insurance (aOR 0.91, 95% CI 0.87–0.94; p<0.001), residence in the highest income quartile (aOR 0.91, 95% CI 0.87–0.94; p<0.001), and treatment at private, non-profit hospitals (aOR 0.89, 95% CI 0.85–0.94; p<0.001).

**Conclusion:** This large nationwide patient database analysis demonstrates that mortality following CPR for IHCA declined before the COVID-19 pandemic but increased during 2020, remaining above pre-pandemic levels through 2022. Independent predictors of mortality reveal socioeconomic disparities in mortality outcomes.

**Question:** How did mortality following cardiopulmonary resuscitation (CPR) for in-hospital cardiac arrest (IHCA) change before, during, and after the COVID-19 pandemic, and what factors were associated with mortality?

**Findings:** In this cross-sectional study of 612,930 weighted hospitalizations from 2016 through 2022, risk-adjusted mortality declined through 2019, increased during the pandemic, peaked in 2021, and remained above pre-pandemic levels in 2022. Higher mortality was independently associated with age, female sex, Black and Hispanic race/ethnicity, and self-pay insurance.

**Meaning:** The findings suggest mortality following CPR for IHCA continues to be elevated after the pandemic peak with certain demographic and socioeconomic groups persistently experiencing poorer outcomes.

## Introduction

In-hospital cardiac arrest (IHCA) remains a major public health burden on inpatient mortality in the United States. Based on the Get with the Guidelines Resuscitation (GWTG-R) database in 2023, 292,000 adults undergo IHCA annually. This statistic has risen from the 211,000 per year prior to 2010. ^1,2^ Survival to discharge after IHCA remains persistently low at 23.6% in 2023, with 79.2% of survivors maintaining a favorable neurological outcome upon discharge.^2^ The American Heart Association (AHA) guidelines for cardiopulmonary resuscitation (CPR) algorithm currently emphasize the importance of early recognition of cardiac arrest, activation of emergency response system, high-quality chest compressions, and defibrillation. ^3^

In the past two decades, studies have demonstrated major improvements in survival after CPR for IHCA, especially since the early 2000s. From 2000 until 2010, risk-adjusted survival to discharge improved from 13.7% to 22.3%.^4^ Analyses of the late 2010s demonstrated mortality after IHCA improved but reached a plateau in the second half of the decade. ^5,6^ These findings emphasize the importance of finding modifiable predictors of mortality to improve outcomes.

In more recent years, National Inpatient Sample (NIS) data for CPR mortality rates post-IHCA revealed a surge in risk-adjusted mortality from 67.4% to 75.4% (2016-2020). ^7^ Limited data exist regarding the post-arrest mortality after the peak of the pandemic. GWTG-R data shows that post-CPR survival improved from 18.8% during the pandemic to 23.6% in 2023, which still is lower than the pre-pandemic survival rate of ∼26.7%.^8^ Survival rates for out-of-hospital cardiac arrest assessed via the US CARES data similarly were below pre-pandemic baseline well into 2022, only showing modest improvements as of 2024. ^8,9^ However, out-of-hospital cardiac arrest (OHCA) outcomes may be influenced by some bystanders’ unwillingness to perform mouth-to-mouth, especially in the pandemic years. ^10^ Regarding OHCA, current literature suggests that there is no significant difference in mortality and survival to hospital admission (after ROSC) between chest compressions only and standard CPR. ^11^ Post-pandemic risk-adjusted mortality trends remain incompletely characterized.

Race-based disparities in IHCA outcomes have been demonstrated with Black patients having lower survival than White patients.^12,13^ Even though mortality rates after CPR were found to improve from 2010 to 2019, this trend was not present in elderly and non-White patients.^14^ It is important to assess the persistence of racial discrepancies in CPR mortality after COVID-19.

Socioeconomic disparities have also been observed, with uninsured patients having worse survival after IHCA.^15^ However, the contribution of insurance status to contemporary IHCA mortality has not been evaluated in recent data.

Prior studies have primarily investigated mortality during the pandemic, so a national analysis in the years following the COVID peak is needed to better understand current trends. We used the HCUP NIS to characterize temporal trends in risk-adjusted in-hospital mortality among adults undergoing CPR for IHCA between 2016 and 2022 and identify independent demographic, hospital, and socioeconomic predictors of mortality. We hypothesized that mortality increased during the COVID-19 pandemic, remained elevated through the early post-pandemic period, and that disparities by race/ethnicity and insurance status would persist after adjustment.

## Methods

### Data Source

The data for this study was derived from National Inpatient Sample (NIS), Healthcare Cost and Utilization Project (HCUP). The NIS database contains weighted discharge data for approximately 35 million hospitalizations annually, representing a 20% stratified sample of discharges from U.S. community hospitals and approximately 98% of the U.S. population. ^16^ Because HCUP NIS data are publicly available and deidentified, this study was exempt from institutional review board approval.

### Study Population

The NIS database years 2016 to 2022 were considered when generating the study population. The NIS database was queried, and the study population was generated, using both International Classification of Diseases, Tenth Revision, Clinical Modification (ICD-10-CM) as well as International Classification of Diseases, Tenth Revision, Procedure Coding System (ICD-10-PCS) codes. Adult patients (over age 20) with cardiac arrest were identified using ICD-10 codes I46.9, I46.0, I46.2, and I46.8. To ensure all patients in the cohort also received either manual or automatic CPR, ICD-10-PCS codes 5A12012, 5A1221Z, and 5A1221J were used. The combination of the diagnosis codes and the ICD-10-PCS code allowed for exclusion of patients that underwent cardiac arrest but received no CPR due to advanced directives or otherwise. Race was determined by the HCUP NIS via hospital billing records.

### Study Outcomes

The primary measured outcome was patient mortality after CPR for IHCA prior to discharge. Analysis of mortality was performed by a using multivariable survey-weighted logistic regression model which was adjusted for age upon admission, sex, race/ethnicity, primary expected payer, median household income quartile of patient’s ZIP code, hospital bed size, hospital status, hospital location, geographic region, and hospital ownership.

### Statistical Analysis

All statistical analyses were conducted using SAS 9.4 (SAS Institute Inc., Cary, NC) and STATA 19 (StataCorp, College Station, TX). Datasets with application of HCUP-provided discharge weights, strata, and cluster variables to account for the complex survey design and generate nationally representative estimates. All reported counts represent weighted estimates of U.S. adult inpatient discharges rather than unique individuals.

Baseline patient demographics, socioeconomic indicators, and hospital-level characteristics were summarized using standard descriptive statistics. Categorical variables were reported as weighted frequencies and percentages, while continuous variables were reported as means with standard deviations (SD) or medians with interquartile ranges (IQR), as appropriate. A complete-case analysis approach was utilized; patients with missing data for primary covariates or outcomes—specifically age, sex, race, length of stay, or in-hospital mortality— were excluded from the final analytical cohort using listwise deletion.

To evaluate independent predictors of post-CPR outcomes, we constructed a multivariable survey-weighted logistic regression model. The primary dependent variable was in-hospital mortality. The model systematically adjusted for patient age at admission, sex, race/ethnicity, primary expected payer, median household income quartile of the patient’s ZIP code, hospital bed size, hospital teaching status/location, geographic region, and hospital ownership control. The independent association of each covariate with the primary outcome was expressed as an adjusted odds ratio (aOR) with a corresponding 95% confidence interval (CI).

To analyze the adjusted temporal trends in mortality across the study period (2016–2022), the admission year was included in the multivariable model as a categorical independent variable. We subsequently employed post-estimation predictive margins to calculate the risk-adjusted predicted probability of mortality for each year. This technique allowed us to estimate the expected mortality rate as if the entire patient cohort were subjected to the specific conditions of a given year, holding all other multivariable covariates constant at their mean distributions. Standard errors and 95% CIs for the predictive margins were calculated utilizing the Delta method. Hypothesis testing was two-tailed, and an alpha level of p < 0.05 was considered statistically significant for all analyses. This study followed the Strengthening the Reporting of Observational Studies in Epidemiology (STROBE) reporting guidelines.

## Results

Between 2016 and 2022, a total of 612,930 weighted hospital discharges for adults (age > 20) meeting the inclusion criteria were identified. The mean age of the cohort was 65.5 years (SD ± 15.2), with a median age of 67 years (IQR: 57–77). The population was predominantly male (58.4%, n = 358,105) and of White race (59.7%, n = 365,845), followed by Black (22.1%, n = 135,420) and Hispanic (11.2%, n = 68,800) patients.

### Temporal Trends in Admissions and Mortality (2016–2022)

Analysis of annual trends revealed a significant fluctuation in admission volumes and clinical outcomes over the study period (p < 0.001). Total annual population estimates remained relatively stable from 2016 to 2019 (ranging from 62,565 to 76,125). However, a marked surge in cases was observed during the pandemic years, peaking in 2021 with 121,565 weighted admissions.

Overall crude inpatient mortality for the study period was 67.8% (n = 415,355). In tandem with the increased admission volume in 2020 and 2021, crude mortality rates exhibited a parallel, statistically significant increase (p < 0.001). Mortality rose from a pre-pandemic baseline of 66.3% in 2016 to 70.2% in 2020, reaching a peak of 71.2% in 2021, before slightly receding to 68.2% in 2022. The median length of stay (LOS) remained stable at approximately 5 days throughout most of the study period, with a transient mean increase in 2021 and 2022 (p < 0.001).

After adjusting for patient demographics, socioeconomic indicators, and hospital characteristics, multivariable logistic regression using predictive margins demonstrated a notable, non-linear pattern in survival outcomes over time (Table 2, Figure 2). Between 2016 and 2019, the risk-adjusted probability of in-hospital mortality following CPR exhibited a gradual, steady decline, reaching its lowest point in 2019 at 63.9% (95% CI: 63.0%–64.8%). However, this favorable trend abruptly reversed at the onset of the COVID-19 pandemic. The adjusted probability of mortality rose sharply to 70.0% (95% CI: 69.2%–70.7%) in 2020 and peaked at 71.3% (95% CI: 70.5%–72.0%) in 2021. By 2022, a partial recovery was observed as the adjusted probability decreased to 68.2% (95% CI: 67.5%–69.0%), though mortality remained substantially elevated above pre-pandemic baseline levels.

**Figure 1:**
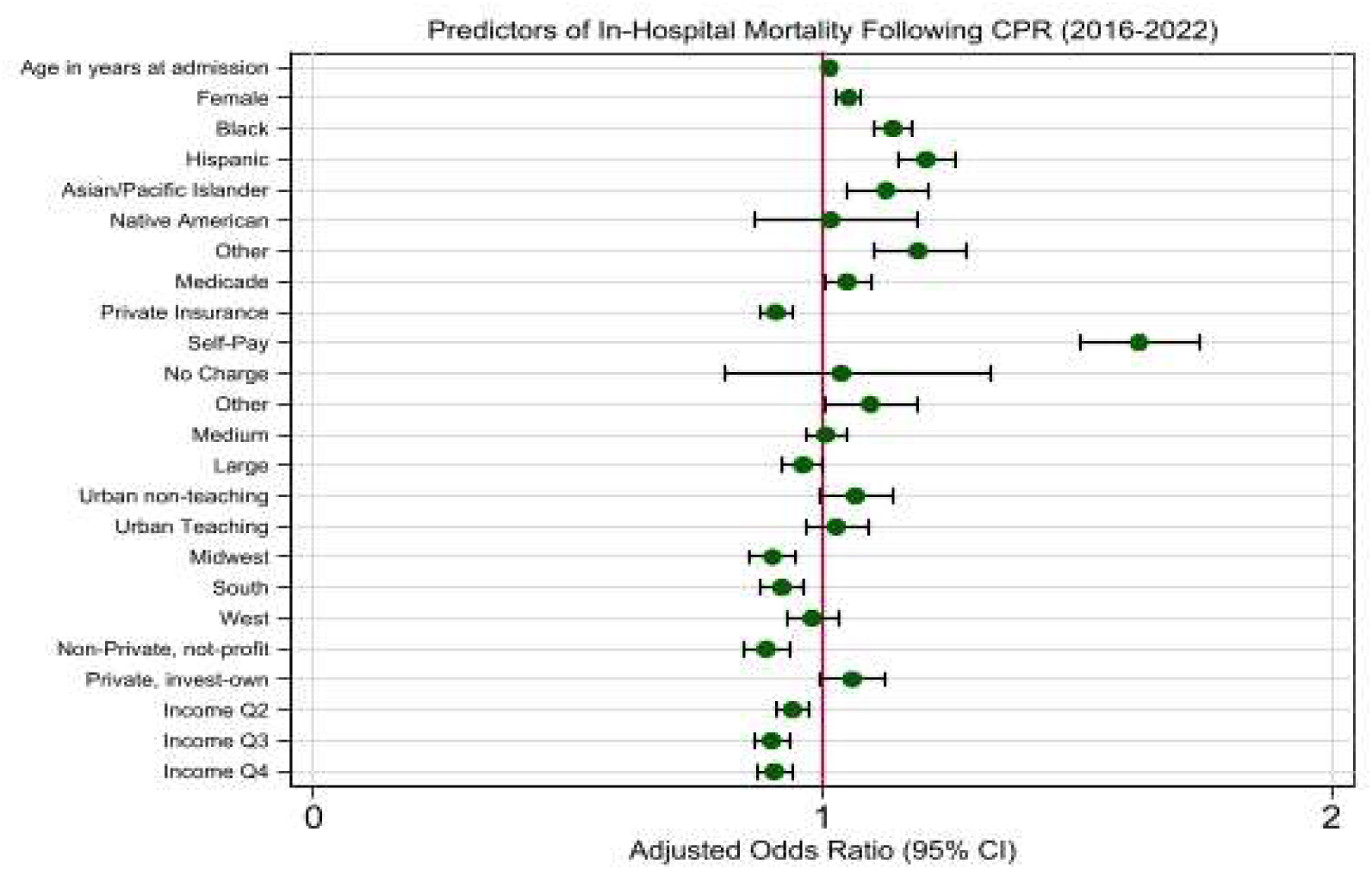
Multivariate Predictors of In-Hospital Mortality. The independent impacts of clinical, demographic, and hospital-level characteristics on post-CPR survival.

**Figure 2:**
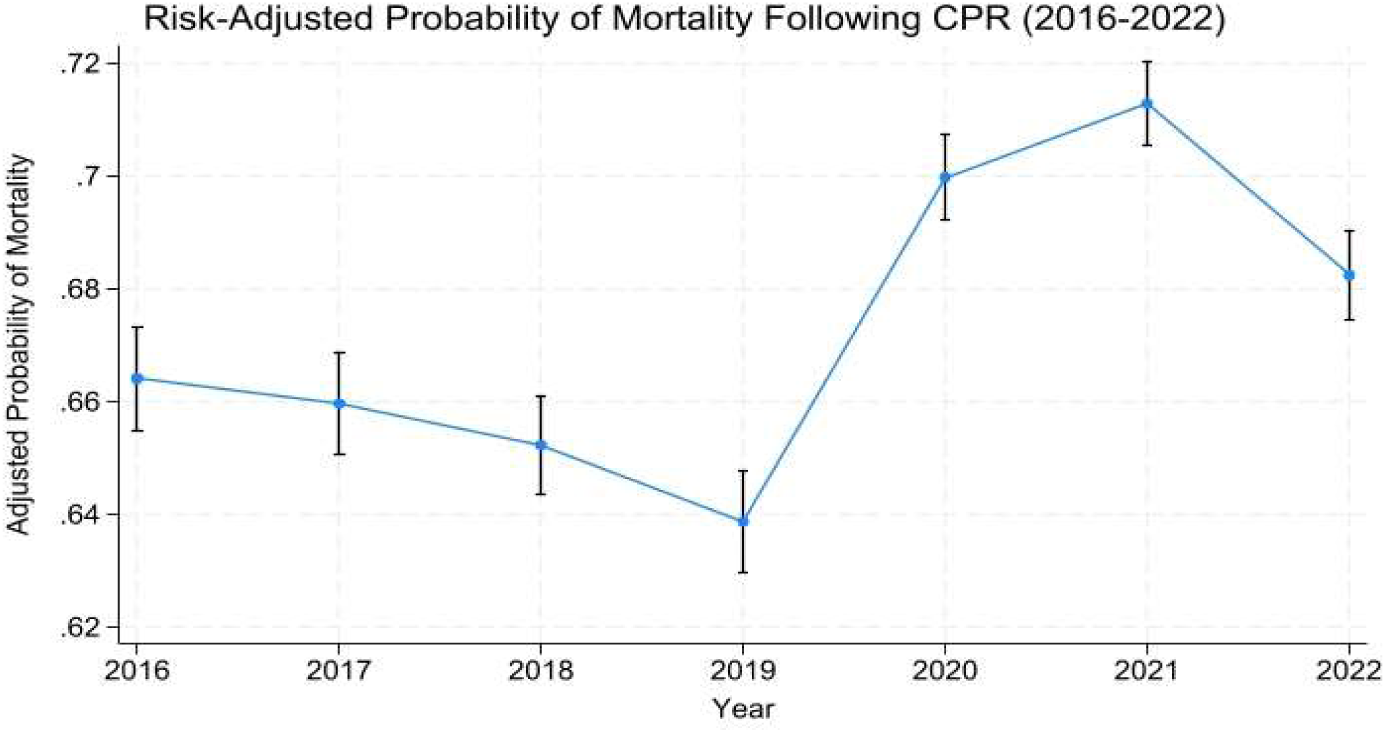
Temporal Trends in Risk-Adjusted Mortality. Risk-adjusted mortality rates after CPR from 2016-2022.

**Table 1:** Demographics of Included Patients. Temporal Trends in Risk-Adjusted Mortality.Length of stay.

|  | **Total** | 2016 | 2017 | 2018 | 2019 | 2020 | 2021 | 2022 | *p-value* |
| --- | --- | --- | --- | --- | --- | --- | --- | --- | --- |
| Population | 612,930 | 62,565 | 72,160 | 75,510 | 76,125 | 98,025 | 121,565 | 106,980 |  |
| Mortality | 415,355<br>(67.8%) | 41,510<br>(66.3%) | 47,570<br>(65.9%) | 49,345<br>(65.3%) | 48,600<br>(63.8%) | 68,785<br>(70.2%) | 86,545<br>(71.2%) | 73,000<br>(68.2%) | <0.001 |
| Age |  |  |  |  |  |  |  |  | <0.001 |
| Mean±SD | 65.5±15.2 | 65.6±15.4 | 65.6±15.3 | 65.6±15.3 | 65.6±15.2 | 65.5±15.1 | 64.8±15.1 | 65.7±15.1 |  |
| Median(IQR) | 67 (57-77) | 67 (57-77) | 67 (57-77) | 67 (57-77) | 67 (57-77) | 67 (57-77) | 67 (56-76) | 67 (57-77) |  |
| LOS |  |  |  |  |  |  |  |  | <0.001 |
| Mean±SD | 9±14 | 9±13 | 9±14 | 9±13 | 9±14 | 9±13 | 10±14 | 10±16 |  |
| Median(IQR) | 5 (1-12) | 4(1-11) | 5 (1-11) | 5 (1-11) | 5 (1-11) | 5 (1-12) | 6 (2-13) | 5 (1-12) |  |
| Gender |  |  |  |  |  |  |  |  | <0.001 |
| Male | 358,105<br>(58.4%) | 35,290<br>(56.4%) | 41,705<br>(57.8%) | 43,385<br>(57.5%) | 44,025<br>(57.8%) | 57,840<br>(59.0%) | 72,680<br>(59.8%) | 63,180<br>(59.1%) |  |
| Female | 254,825<br>(41.6%) | 27,275<br>(43.6%) | 30,455<br>(42.2%) | 32,125<br>(42.5%) | 32,100<br>(42.2%) | 40,185<br>(41.0%) | 48,885<br>(40.2%) | 43,800<br>(40.9%) |  |
| Race |  |  |  |  |  |  |  |  | <0.001 |
| White | 365,845<br>(59.7%) | 39,050<br>(62.4%) | 44,480<br>(61.6%) | 46,020<br>(60.9%) | 46,850<br>(61.5%) | 55,030<br>(56.1%) | 69,990<br>(57.6%) | 64,425<br>(60.2%) |  |
| Black | 135,420<br>(22.1%) | 13,700<br>(21.9%) | 15,460<br>(21.4%) | 16,195<br>(21.4%) | 16,440<br>(21.6%) | 22,820<br>(23.3%) | 27,690<br>(22.8%) | 23,115<br>(21.6%) |  |
| Hispanic | 68,800 (11.2%) | 5,820 (9.3%) | 7,300 (10.1%) | 8,205 (10.9%) | 7,735 (10.2%) | 12,960<br>(13.2%) | 14,995<br>(12.3%) | 11,785<br>(11.0%) |  |
| Asian/Pacific Islander | 19,665 (3.2%) | 1,800 (2.9%) | 2,200 (3.0%) | 2,300 (3.0%) | 2,390 (3.1%) | 3,190 (3.3%) | 4,190 (3.4%) | 3,595 (3.4%) |  |
| Native American | 3,870 (0.6%) | 330 (0.5%) | 475 (0.7%) | 415 (0.5%) | 515 (0.7%) | 620 (0.6%) | 795 (0.7%) | 720 (0.7%) |  |
| Others | 19,330 (3.2%) | 1,865 (3.0%) | 2,245 (3.1%) | 2,375 (3.1%) | 2,195 (2.9%) | 3,405 (3.5%) | 3,905 (3.2%) | 3,340 (3.1%) |  |
| Primary Payer |  |  |  |  |  |  |  |  | <0.001 |
| Medicare | 369,305<br>(60.3%) | 38,785<br>(62.0%) | 45,015<br>(62.5%) | 47,185<br>(62.6%) | 46,705<br>(61.4%) | 58,380<br>(59.6%) | 69,455<br>(57.2%) | 63,780<br>(59.7%) |  |
| Medicaid | 88,090 (14.4%) | 8,190 (13.1%) | 9,585 (13.3%) | 9,990 (13.2%) | 10,250<br>(13.5%) | 14,575<br>(14.9%) | 19,205<br>(15.8%) | 16,295<br>(15.2%) |  |
| Private including HMO | 109,595<br>(17.9%) | 11,125<br>(17.8%) | 12,615<br>(17.5%) | 12,950<br>(17.2%) | 13,320<br>(17.5%) | 17,565<br>(17.9%) | 23,050<br>(19.0%) | 18,970<br>(17.8%) |  |
| Self-Pay | 26,660 (4.4%) | 2,585 (4.1%) | 3,075 (4.3%) | 3,340 (4.4%) | 3,635 (4.8%) | 4,290 (4.4%) | 5,435 (4.5%) | 4,300 (4.0%) |  |
| No Charge | 1,545 (0.3%) | 170 (0.3%) | 105 (0.1%) | 175 (0.2%) | 205 (0.3%) | 270 (0.3%) | 370 (0.3%) | 250 (0.2%) |  |
| Other | 17,050 (2.8%) | 1,665 (2.7%) | 1,675 (2.3%) | 1,795 (2.4%) | 1,925 (2.5%) | 2,850 (2.9%) | 3,865 (3.2%) | 3,275 (3.1%) |  |
| Hospital Bed size |  |  |  |  |  |  |  |  | <0.001 |
| Small | 112,900<br>(18.4%) | 9,215 (14.7%) | 10,575<br>(14.7%) | 12,785<br>(16.9%) | 14,335<br>(18.8%) | 20,085<br>(20.5%) | 25,220<br>(20.7%) | 20,685<br>(19.3%) |  |
| Medium | 179,780<br>(29.3%) | 17,655<br>(28.2%) | 21,915<br>(30.4%) | 22,845<br>(30.3%) | 22,375<br>(29.4%) | 28,320<br>(28.9%) | 34,585<br>(28.4%) | 32,085<br>(30.0%) |  |
| Large | 320,250<br>(52.2%) | 35,695<br>(57.1%) | 39,670<br>(55.0%) | 39,880<br>(52.8%) | 39,415<br>(51.8%) | 49,620<br>(50.6%) | 61,760<br>(50.8%) | 54,210<br>(50.7%) |  |
| Hospital Teaching/Location |  |  |  |  |  |  |  |  | <0.001 |
| Rural | 33,940 (5.5%) | 3,560 (5.7%) | 3,695 (5.1%) | 3,820 (5.1%) | 3,900 (5.1%) | 5,070 (5.2%) | 7,700 (6.3%) | 6,195 (5.8%) |  |
| Urban non-teaching | 121,370<br>(19.8%) | 16,560<br>(26.5%) | 16,085<br>(22.3%) | 15,520<br>(20.6%) | 13,570<br>(17.8%) | 18,695<br>(19.1%) | 22,910<br>(18.8%) | 18,030<br>(16.9%) |  |
| Urban Teaching | 457,620<br>(74.7%) | 42,445<br>(67.8%) | 52,380<br>(72.6%) | 56,170<br>(74.4%) | 58,655<br>(77.1%) | 74,260<br>(75.8%) | 90,955<br>(74.8%) | 82,755<br>(77.4%) |  |
| Hospital Region |  |  |  |  |  |  |  |  | 0.98 |
| Northeast | 88,680 (14.5%) | 9,700 (15.5%) | 10,850<br>(15.0%) | 10,825<br>(14.3%) | 10,895<br>(14.3%) | 14,275<br>(14.6%) | 16,600<br>(13.7%) | 15,535<br>(14.5%) |  |
| Midwest | 127,620<br>(20.8%) | 13,120<br>(21.0%) | 15,575<br>(21.6%) | 16,595<br>(22.0%) | 16,275<br>(21.4%) | 19,440<br>(19.8%) | 23,820<br>(19.6%) | 22,795<br>(21.3%) |  |
| South | 268,410<br>(43.8%) | 27,565<br>(44.1%) | 31,050<br>(43.0%) | 32,660<br>(43.3%) | 33,205<br>(43.6%) | 43,425<br>(44.3%) | 54,825<br>(45.1%) | 45,680<br>(42.7%) |  |
| West | 128,220<br>(20.9%) | 12,180<br>(19.5%) | 14,685<br>(20.4%) | 15,430<br>(20.4%) | 15,750<br>(20.7%) | 20,885<br>(21.3%) | 26,320<br>(21.7%) | 22,970<br>(21.5%) |  |
| Control/ownership of hospital |  |  |  |  |  |  |  |  | 0.94 |
| Government, nonfederal | 69,785 (11.4%) | 7,225 (11.5%) | 8,475 (11.7%) | 8,695 (11.5%) | 8,835 (11.6%) | 11,365<br>(11.6%) | 13,515<br>(11.1%) | 11,675<br>(10.9%) |  |
| Private, not-profit | 446,300<br>(72.8%) | 45,475<br>(72.7%) | 52,375<br>(72.6%) | 55,105<br>(73.0%) | 55,480<br>(72.9%) | 69,770<br>(71.2%) | 88,090<br>(72.5%) | 80,005<br>(74.8%) |  |
| Private, invest-own | 96,845 (15.8%) | 9,865 (15.8%) | 11,310<br>(15.7%) | 11,710<br>(15.5%) | 11,810<br>(15.5%) | 16,890<br>(17.2%) | 19,960<br>(16.4%) | 15,300<br>(14.3%) |  |
| Median Household Income |  |  |  |  |  |  |  |  | 0.48 |
| Quartile 1 | 206,490<br>(34.3%) | 21,500<br>(35.0%) | 24,665<br>(34.8%) | 24,875<br>(33.6%) | 25,190<br>(33.8%) | 34,205<br>(35.5%) | 41,335<br>(34.7%) | 34,720<br>(33.0%) |  |
| Quartile 2 | 153,480<br>(25.5%) | 15,230<br>(24.8%) | 18,320<br>(25.9%) | 19,285<br>(26.1%) | 18,665<br>(25.0%) | 24,915<br>(25.9%) | 29,890<br>(25.1%) | 27,175<br>(25.8%) |  |
| Quartile 3 | 134,845<br>(22.4%) | 13,605<br>(22.2%) | 15,520<br>(21.9%) | 16,245<br>(22.0%) | 17,260<br>(23.1%) | 20,915<br>(21.7%) | 26,815<br>(22.5%) | 24,485<br>(23.3%) |  |
| Quartile 4 | 106,640<br>(17.7%) | 11,010<br>(17.9%) | 12,340<br>(17.4%) | 13,560<br>(18.3%) | 13,505<br>(18.1%) | 16,240<br>(16.9%) | 21,225<br>(17.8%) | 18,760<br>(17.8%) |  |

**Table 2:** Adjusted Predicted Probability of Mortality Following CPR (2016-2022). Abbreviations: REF, reference.

| Year | Adjusted<br>Probability<br>(%) | 95% CI |
| --- | --- | --- |
| 2016 | 66.4 | 65.5 – 67.3 |
| 2017 | 66 | 65.1 – 66.9 |
| 2018 | 65.2 | 64.4 – 66.1 |
| 2019 | 63.9 | 63.0 – 64.8 |
| 2020 | 70 | 69.2 – 70.7 |
| 2021 | 71.3 | 70.5 – 72.0 |
| 2022 | 68.2 | 67.5 – 69.0 |

### Baseline Patient and Hospital Characteristics

Regarding socioeconomic and insurance status, the majority of the cohort was covered by Medicare (60.3%, n = 369,305), with private insurance (17.9%, n = 109,595) and Medicaid (14.4%, n = 88,090) representing the next most common primary payers. Patients falling into the lowest median household income quartile (Quartile 1) represented the largest income subgroup (34.3%, n = 206,490), though temporal variations across income quartiles throughout the study period were not statistically significant (p = 0.48).

The majority of patients were treated at large hospital facilities (52.2%, n = 320,250) and within Urban Teaching institutions (74.7%, n = 457,620). Geographically, the highest proportion of admissions occurred in the South (43.8%, n = 268,410). Most facilities were classified as private, not-for-profit hospitals (72.8%, n = 446,300). There were no significant temporal shifts observed regarding the hospital region (p = 0.98) or the control/ownership of the treating hospitals (p = 0.94) over the seven-year span. The independent impacts of clinical, demographic, and hospital-level characteristics on post-CPR survival are illustrated in Figure 1.

### Multivariate Analysis of Predictors of In-Hospital Mortality

In the multivariable survey-weighted logistic regression model (Table 3), the year of admission remained a significant independent predictor of mortality. Compared to the 2016 baseline, admission in 2019 was associated with significantly lower odds of mortality (aOR 0.89, 95% CI: 0.84–0.95; p<0.001). Conversely, the pandemic years were associated with the highest likelihood of death, with admissions in 2020 (aOR 1.18, 95% CI: 1.12–1.25; p<0.001) and 2021 (aOR 1.26, 95% CI: 1.19–1.33; p<0.001) demonstrating significantly increased mortality odds.

**Table 3:** Multivariate Predictors of In-Hospital Mortality Among Hospitalized Adults with Cardiac Arrest Undergoing CPR. Abbreviations: REF, reference; invest-own, investor owned (for-profit) hospital.

| Variable | Adjusted OR | 95% CI | p-value |
| --- | --- | --- | --- |
| Year (per year increase) |  |  |  |
| 2016 | REF |  |  |
| 2017 | 0.98 | 0.92-1.04 | 0.493 |
| 2018 | 0.95 | 0.90-1.00 | 0.066 |
| 2019 | 0.89 | 0.84-0.95 | <0.001 |
| 2020 | 1.18 | 1.12-1.25 | <0.001 |
| 2021 | 1.26 | 1.19-1.33 | <0.001 |
| 2022 | 1.09 | 1.03-1.15 | 0.003 |
| Age (per year) | 1.01 | 1.01-1.01 | <0.001 |
| Gender |  |  |  |
| Male | REF |  |  |
| Female | 1.05 | 1.02-1.08 | <0.001 |
| Race |  |  |  |
| White | REF |  |  |
| Black | 1.14 | 1.10-1.18 | <0.001 |
| Hispanic | 1.2 | 1.15-1.26 | <0.001 |
| Asian/Pacific Islander | 1.12 | 1.05-1.21 | 0.002 |
| Native American | 1.01 | 0.87-1.19 | 0.861 |
| Others | 1.19 | 1.10-1.28 | <0.001 |
| Primary Payer |  |  |  |
| Medicare | REF |  |  |
| Medicaid | 1.05 | 1.00-1.09 | 0.032 |
| Private including HMO | 0.91 | 0.87-0.94 | <0.001 |
| Self-Pay | 1.62 | 1.51-1.74 | <0.001 |
| No Charge | 1.04 | 0.81-1.33 | 0.784 |
| Other | 1.09 | 1.01-1.19 | 0.033 |
| Hospital Bed size |  |  |  |
| Small | REF |  |  |
| Medium | 1.01 | 0.97-1.05 | 0.756 |
| Large | 0.96 | 0.92-1.00 | 0.05 |
| Hospital Teaching/Location |  |  |  |
| Rural | REF |  |  |
| Urban non-teaching | 1.07 | 1.00-1.14 | 0.061 |
| Urban Teaching | 1.03 | 0.97-1.09 | 0.402 |
| Hospital Region |  |  |  |
| Northeast | REF |  |  |
| Midwest | 0.9 | 0.86-0.95 | <0.001 |
| South | 0.92 | 0.88-0.96 | <0.001 |
| West | 0.98 | 0.93-1.03 | 0.45 |
| Control/ownership of hospital |  |  |  |
| Government, nonfederal | REF |  |  |
| Private, not-profit | 0.89 | 0.85-0.94 | <0.001 |
| Private, invest-own | 1.06 | 1.00-1.12 | 0.069 |
| Median Household Income |  |  |  |
| Quartile 1 | REF |  |  |
| Quartile 2 | 0.94 | 0.91-0.97 | <0.001 |
| Quartile 3 | 0.9 | 0.87-0.93 | <0.001 |
| Quartile 4 | 0.91 | 0.87-0.94 | <0.001 |

Several patient-level and hospital-level factors were also independently associated with increased mortality. Increasing age was associated with a slight, incremental increase in mortality risk (aOR 1.01 per year; p<0.001), as was female sex (aOR 1.05, 95% CI: 1.02–1.08; p<0.001) compared to male sex. Furthermore, significant racial disparities were observed; compared to White patients, Black (aOR 1.14, 95% CI: 1.10–1.18; p<0.001), Hispanic (aOR 1.20, 95% CI: 1.15–1.26; p<0.001), Asian/Pacific Islander (aOR 1.12, 95% CI: 1.05–1.21; p=0.002), and Other race (aOR [1.19], 95% CI: [1.10–1.28]; p<0.001) patients all faced significantly higher odds of death following CPR.

Socioeconomic and healthcare access indicators also strongly predicted outcomes. Self-pay status was associated with the highest increased odds of mortality compared to Medicare (aOR 1.62, 95% CI: 1.51–1.74; p<0.001). Conversely, private insurance (aOR 0.91, 95% CI: 0.87–0.94; p<0.001) and admission to a private, non-profit hospital (aOR 0.89, 95% CI: 0.85– 0.94; p<0.001) were protective. Higher neighborhood income quartiles were associated with a stepwise decrease in mortality odds, with patients in the highest income quartile (Quartile 4) demonstrating a 9% reduction in the odds of death (aOR 0.91, 95% CI: 0.87–0.94; p<0.001) compared to those in the lowest quartile.

## Discussion

This is the most recent multi-year HCUP-NIS analysis, characterizing risk-adjusted mortality trends spanning the pre-pandemic, pandemic, and early post-pandemic periods until 2022 among patients undergoing CPR for IHCA. The nonlinear trend for risk adjusted mortality displays a downward trend from 2016-2019, followed by a sharp rise during the onset of COVID-19, which did not return to baseline as of 2022.

Infection during the pandemic has been shown to inflate the mortality rates post CPR, with Swedish registry data demonstrating 2.3-fold higher 30-day mortality in COVID-positive IHCA compared to COVID-negative arrests.^17^ In the United States, NIS data has similarly demonstrated COVID-19 patients who arrested had 3.9-fold higher adjusted odds of mortality than patients without COVID-19.^7^ The relationship between COVID-19 infection and the notable mortality increase post-CPR in 2020 suggests COVID-19 infection may have been a contributing factor regarding the rise in mortality observed in 2020.

Besides the pathophysiological effects of COVID-19, evidence suggests that systemic hospital-level strain also contributed to worse outcomes during the pandemic. Get With the Guidelines–Resuscitation (GWTG-R) registry data from 61,586 IHCAs demonstrated earlier termination of CPR among patients with cardiac arrest during the initial surge of the pandemic (March–May 2020). Lower post-CPR survival persisted even after excluding patients with COVID-19, suggesting that factors beyond SARS-CoV-2 infection, including changes in resuscitation practices or reduced quality of care, may have contributed to poorer outcomes.^18^ It is plausible that unprecedented strain on hospital staff, ICU capacity, and healthcare resources negatively affected the delivery of resuscitation care, ultimately contributing to the increased mortality observed during the pandemic.

There is also evidence that patients experiencing IHCA may have entered hospitalization with more advanced or poorly controlled disease because of delayed healthcare utilization during the pandemic. Retrospective models have estimated that national ICU occupancy exceeding 75% was associated with approximately 12,000 excess deaths over a two-week period. Additionally, emergency department visits for myocardial infarction, diabetic emergencies, and cerebrovascular accidents declined substantially in 2020, suggesting that many patients delayed seeking care until their conditions had progressed.^19,20^ Such delays may have resulted in patients experiencing cardiac arrest with greater underlying physiologic compromise, thereby reducing the likelihood of successful resuscitation and survival.

Prior evidence is consistent in showing that post-CPR survival has not improved from pre-pandemic baseline.^8^ Persistent excess mortality after the acute pandemic phase suggests that the pandemic’s effects extended beyond acute SARS-CoV-2 infection alone. From 2018-2021, over 1 billion medication records from the UK found 491,306 fewer individuals started antihypertensive medication than expected between March 2020 and July 2021. Lipid-lowering medication usage also decreased in the first half of 2021 compared to 2019. These untreated patients were predicted to experience 13,662 future cardiovascular disease events over their life including 2,281 myocardial infarctions and 3,474 strokes.^21^ Deferred preventative care and treatment during the peak of the pandemic may have contributed to the post-CPR mortality rates staying above the pre-pandemic baseline.^18,22^ Lack of care during the pandemic may be due to postponed hospitalizations for acute events, limited access to medications and diagnostic testing, increased sedentary behavior, and general worsening of cardiometabolic risk factor control.^23^

Our analysis revealed that Black, Hispanic, and Asian/Pacific Islander patients were significant predictors of IHCA mortality post-CPR. Demographically, Black male and female patients from 2016-2020 have been historically found to have higher odds of mortality from VT/VF arrests and PEA/asystole arrests when compared to White male. Notably, Black and Hispanic male patients also had significantly lower odds of receiving PCI and CABG when compared to White males after cardiac arrest.^12^ Such discrepancies in cardiovascular interventions suggest a possible contribution to persistent racial disparity in mortality after IHCA and CPR. Especially given the strain the COVID-19 pandemic placed on ICUs and hospital wards across the country, it is plausible that existing racial disparities in the likelihood of receiving procedures may have persisted or even widened.

The association between sex and mortality following IHCA have demonstrated inconsistent sex-based differences across studies. Our adjusted model found that female sex was independently associated with higher odds of in-hospital mortality after IHCA and CPR, consistent with recent national data.^24^ In contrast, a prior meta-analysis reported lower survival among males following cardiac arrest.^25^ These discrepancies may partially reflect differences in study populations and age distributions. Women of childbearing age have demonstrated better outcomes than age-matched men, whereas this evident survival advantage appears to diminish with advancing age.^1^ The survival advantage observed among women of childbearing age has been hypothesized to reflect the protective effects of endogenous sex hormones, although the underlying mechanisms are not fully understood.^26^

Advanced age is a well-established predictor of mortality following IHCA, although it is generally less prognostic than clinical factors such as initial arrest rhythm.^1,27^ Our analysis demonstrated that each additional year of age was associated with a 1% increase in the adjusted odds of mortality.

Existing literature is mixed for the impact of the hospital type on mortality rates. From 2008-2012, lower mortality rates after cardiac arrest were found in teaching hospitals as against non-teaching hospitals, although this difference disappeared after adjusting for procedures suggesting higher utilization of interventions at teaching hospitals.^28^ Other studies have shown urban and large teaching hospitals have higher mortality, possibly biased by high-acuity patients being transferred to such hospitals with more capabilities and resources.^29^ The results of our analysis demonstrate similar results, in that urban hospitals have nonsignificant higher likelihood of mortality post-arrest. These findings suggest that after adjustment for patient and hospital characteristics, hospital teaching status and urban location may contribute less to mortality than patient-level clinical factors.

The relationship between health insurance status and mortality outcomes after CPR has been studied, although there remain some gaps which have been addressed by our study. An NIS study investigating out-of-hospital VF arrest from 2003 to 2014 found that uninsured status was independently associated with higher in-hospital mortality and lower utilization of implantable cardioverter-defibrillators (ICDs).^15^ The poorer outcomes for uninsured patients might involve the lack of insurance coverage for ICDs as well as worse baseline cardiovascular health with less regular preventative care. Prior studies have demonstrated poorer outcomes among uninsured patients following cardiac arrest.^15^ We found self-pay status to be independently associated with mortality, potentially reflecting disparities in access to preventive care and chronic disease management. Although Pancholy’s study was solely focused on out-of-hospital VF arrest, an overlap can be drawn between uninsured status and our study’s self-pay patient population who were predicted to have higher risk of mortality after IHCA and CPR.^15^ Our findings align with prior literature, revealing that self-pay insurance status is independently associated with higher risk of mortality, possibly due to limited access to care which results in them having worse outcomes after IHCA.

As our study utilizes the HCUP-NIS database, there are some inherent limitations to be acknowledged. First, the administrative nature of the database precludes assessment of important resuscitation-specific variables, such as CPR quality metrics, duration of resuscitation, return of spontaneous circulation, neurological status, and initial arrest rhythm. Although prior studies have suggested that CPR quality declined during and beyond the COVID-19 pandemic, these data are unavailable within the NIS, preventing direct evaluation of their contribution to the observed mortality trends.^18,30^ Additionally, the database lacks information regarding outpatient medication use, longitudinal healthcare utilization, and disease management before hospitalization, limiting our ability to account for baseline cardiovascular risk and chronic disease control. As such, residual confounding from unmeasured clinical variables may be present despite multivariable adjustment. These limitations are particularly relevant when interpreting the association between self-pay insurance status and mortality. The NIS does not capture factors such as access to preventive care, medication adherence, or severity of underlying disease before hospitalization. While self-pay status remained independently associated with increased mortality, the mechanisms underlying this relationship cannot be determined from administrative data alone and warrant further investigation using datasets with greater clinical granularity.

### Conclusion

This nationwide HCUP-NIS analysis demonstrates that the favorable decline in risk-adjusted mortality following IHCA and CPR observed before the COVID-19 pandemic was interrupted by a marked increase in mortality during 2020, with outcomes remaining above pre-pandemic levels through 2022. These findings characterize national mortality trends following CPR during and after the pandemic while identifying persistent demographic and socioeconomic disparities in survival rates. Future studies incorporating detailed clinical and resuscitation data could identify modifiable factors underlying these trends, potentially allowing for improvements in outcomes for patients experiencing IHCA.

## Data Availability

The NIS data are available from the Healthcare Cost and Utilization Project (HCUP) after completion of the required Data Use Agreement. The data cannot be publicly shared by the authors due to the required DUA.

## Acknowledgement

All authors declare no conflict of interest. No funding was utilized for this project.

All authors contributed to the manuscript and take responsibility for its accuracy.

Mehrtash Hashemzadeh had full access to all the data in the study and was responsible for the data analysis.

Study concept and design: All authors.

Acquisition, analysis, or interpretation of data: All authors.

Drafting of the manuscript: Nedunuri, Rahul

Critical revision of the manuscript for important intellectual content: All authors.

Statistical analysis: Hashemzadeh, Mehrtash

Administrative, technical, or material support: Hashemzadeh, Mehrtash

Study supervision: Movahed, Mohammad Reza

